# Modelling the Potential Role of Media Campaigns on the Control of Listeriosis

**DOI:** 10.1101/2020.12.22.20248698

**Authors:** C. W. Chukwu, F. Nyabadza, Fatmawati

**Affiliations:** Department of Mathematics and Applied Mathematics, University of Johannesburg, Auckland Park Kingsway Campus, PO Box 524, 2006, Johannesburg, South Africa; Department of Mathematics, Faculty of Science and Technology, Universitas Airlangga, Surabaya 60115, Indonesia

**Keywords:** Equilibria, Listeria, Numerical Simulations, Food contamination threshold

## Abstract

Human Listeria infection is a food-borne disease caused by the consumption of contaminated food products by the bacterial pathogen, Listeria. In this paper, we propose a mathematical model to analyze the impact of media campaigns on the spread and control of Listeriosis. The model exhibited three equilibria namely; disease-free, Listeria-free and endemic equilibria. The food contamination threshold (ℛ_*f*_) is determined and the local stability analyses of the model discussed. Sensitivity analysis is done to determine the model parameters that most affect the severity of the disease. Numerical simulations were carried out to assess the role of media campaigns on the Listeriosis spread. The results show that; an increase in the intensity of the media awareness campaigns, the removal rate of contaminated food products, a decrease in the contact rate of Listeria by humans results in fewer humans getting infected, thus leading to the disease eradication. An increase in the depletion of media awareness campaigns results in more humans being infected with Listeriosis. These findings may significantly impact policy and decision making in the control of Listeriosis disease.

## 1 Introduction

Listeriosis is a serious and severe food-borne disease that affects the human population globally. The disease is caused by a bacteria called *Listeria monocytogenes* which exists in the environment as its primary host (soil, water, ready-to-eat (RTE) foods and contaminated food products) [1,2]. Human beings contract Listeriosis through the ingestion of contaminated RTE food products such as cantaloupes, meat, Ricotta Salata cheese, vegetables, polony, bean sprouts, ham, or directly from the environment [3, 4]. The epidemiology of Listeriosis is clearly articulated in [1, 4, 12].

Before the 2017 outbreak in South Africa, an average of 60 to 80 confirmed Listeriosis disease cases were recorded annually (i.e., approximately 1 case per week). The recent outbreak in South Africa, which occurred from 1 January 2017 to 17 July 2018, had 1 060 confirmed cases, with 216 (26.8%) deaths. This was the world’s largest ever documented Listeriosis outbreak [6]. The source of the disease was traced to be contaminated RTE processed meat products. Also, Listeriosis outbreaks resulting from human consumption of different kinds of contaminated RTE food products occur commonly in the United States of America, Canada and Europe [3].

Media campaigns and media-driven awareness programs such as print media, social media, internet, television, radio and advertisements play an essential role in the disseminating information about the spread of infectious disease outbreaks [7]. In particular, they influence individuals and the public health care systems’ behavior in the implementation of control strategies to contain the spread of diseases. Dissemination of information educates people and helps them take precautionary measures such as; practising better hygiene; factory workers wearing clean gloves to avoid cross-contamination of food products during food production/manufacturing.

In recent times, researchers have used mathematical models to describe the effect and impact of media campaigns or several awareness programs on the dynamics of infectious such as Ebola [8], Human Immunodeficiency Virus [13], Human Influenza [14] e.t.c. Misra *et al*. [15], used a non-linear mathematical model which assumed that due to awareness programs by media, once the population becomes aware of the disease spread, they avoid contacts with the infectives and therefore form a new class of individuals called the aware class, who become susceptible again if their awareness wanes over time. The model analysis revealed that the number of infectives decreases with an increase in media campaigns. Kaur [16], extended the work by Misra by assuming that aware susceptibles do not lose awareness but can also interact with infected individuals and get infected, albeit at a lower rate. Their study suggested that with the increase in the rate of implementation of awareness programs via media, there is a subsequent decline in the number of infected in any targeted population under consideration. Authors in [8] used a mathematical model to describe the transmission dynamics of Ebola in the presence of asymptomatic cases and the impact of media campaigns on the disease transmission was represented by a linearly decreasing function. Their results showed that messages sent through media have a more significant effect on reducing Ebola cases if they are more effective and spaced out. An *SIRS* model was proposed in [17] to investigate the impact of awareness programs by considering private and public awareness, which reduces the contact rate between unaware and aware population and the effect/impact of public information campaigns on disease prevalence. It was shown that both private and public awareness could reduce the size of epidemic outbreaks. A smoking cessation model with media campaigns was presented [18]. The results showed that the reproduction number was suppressed when media campaigns that focus on smoking cessation were increased. Thus, spreading information to encourage smokers to quit smoking was an effective intervention. According to [19], an *SIRS* model was used to analyze the role of information and limited optimal treatment on disease prevalence. The model considered the growth rate of information proportional to a saturated function of infected individuals. The results from the mathematical analysis showed that the combined effects of information and treatment are more effective and economical in the control of the infection. Exponential functions have also been used to model the impact of media awareness campaigns on people’s behaviour, which affects the evolution of infectious diseases [20, 21]. In particular, the effects of Twitter messages on reducing the transmission rate of the influenza virus was studied in [21]. The result revealed that Twitter messages had a substantial influence on the dynamics of influenza disease spread.

To date, there are very few mathematical models on the dynamics of Listeriosis (see, for instance, [9–12]), let alone those investigating the potential role of media awareness campaigns on the dynamics of Listeriosis. This paper is motivated by the work done in [15]. We formulate a mathematical model to study the impact and effects of media campaigns on the dynamics of Listeriosis disease resulting from the consumption of contaminated RTE in the human population. We describe the model in detail in the following section.

The outline of this paper is as follows; Section 1 introduces the research paper followed by the model described in Section 2. The model basic properties and analyses are presented in Section 3. Numerical simulations were done and presented in Section 4. Section 5 concludes the paper.

## 2 Model description

The human population is divided into four sub-classes, viz: Susceptibles *S*_*h*_(*t*), aware susceptibles *S*_*a*_(*t*), the infected *I*_*h*_(*t*) and the recovered *R*_*h*_(*t*). Individuals are recruited at a rate proportional to the size of the human population *N* (*t*) where

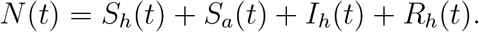

The recruitment rate is given by *µ*_*h*_*N* (*t*) where *µ*_*h*_ is natural birth/mortality rate. Upon infection with Listeria from contaminated food, the susceptibles move into the infectious class *I*_*h*_(*t*) with a force of infection *λ*_*h*_(*t*), where 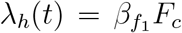 with 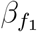 being the rate at which humans gets infected and *F*_*c*_(*t*) the contaminated food products. Here, *λ*_*h*_(*t*) describes the force of infection by the consumption of contaminated food products. The susceptible individuals can also move to the awareness class at a rate *ρ* as a result of the interaction with the media campaigns. We assume that media campaigns wane over time and the aware individuals can revert to being susceptible again at a rate *ω*_1_. The infected individuals recover at a rate *γ* with immunity after treatment. These individuals who recover after some time can also lose their immunity and become susceptible again at a rate *δ*_*h*_. We assume a constant human population *N* (*t*), which consists of individuals who do not work in the factory over the modelling time. Further, we assume that aware individuals cannot be infected as their awareness protects them from contracting the disease. Given that the bacteria survive even at 4°*C*, it can die or grow in its host or the environment at significantly low temperatures. Let *r*_*l*_ and *ξ* denote the growth and removal rate of the of Listeria, *L*(*t*). Our model assumes a logistic growth of Listeria with carrying capacity *K*_*L*_. The non-contaminated food products *F*_*n*_(*t*) can be contaminated as a result of interaction with the bacteria from the environment that comes, via the workers, exchange of gloves or utensils during food manufacturing and also through the contact with contaminated food *F*_*c*_(*t*) with a force of infection *λ*_*f*_ (*t*), where

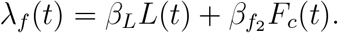

The parameters *β*_*L*_ and 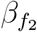 are the contact rate of Listeria and the contamination rate of non-contaminated food by contaminated food products, respectively. The contaminated food products are then responsible for transmitting Listeriosis disease to the human population through ingestion of the contaminated food products. The total amount of food products, *F* (*t*), at any given time is given by

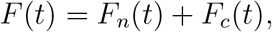

where *µ*_*f*_ is the rate of removal of food products through consumption. Let *M*_*a*_ be the cumulative density of media campaigns with maximum intensity, *M*, at which media awareness campaigns are implemented, *π*_0_ the rate of implementation of the media awareness campaigns and *µ*_0_ the rate of depletion of media awareness. The above model descriptions and Figure 1 gives the following systems of non-linear ordinary differential equations:

**Figure 1:**
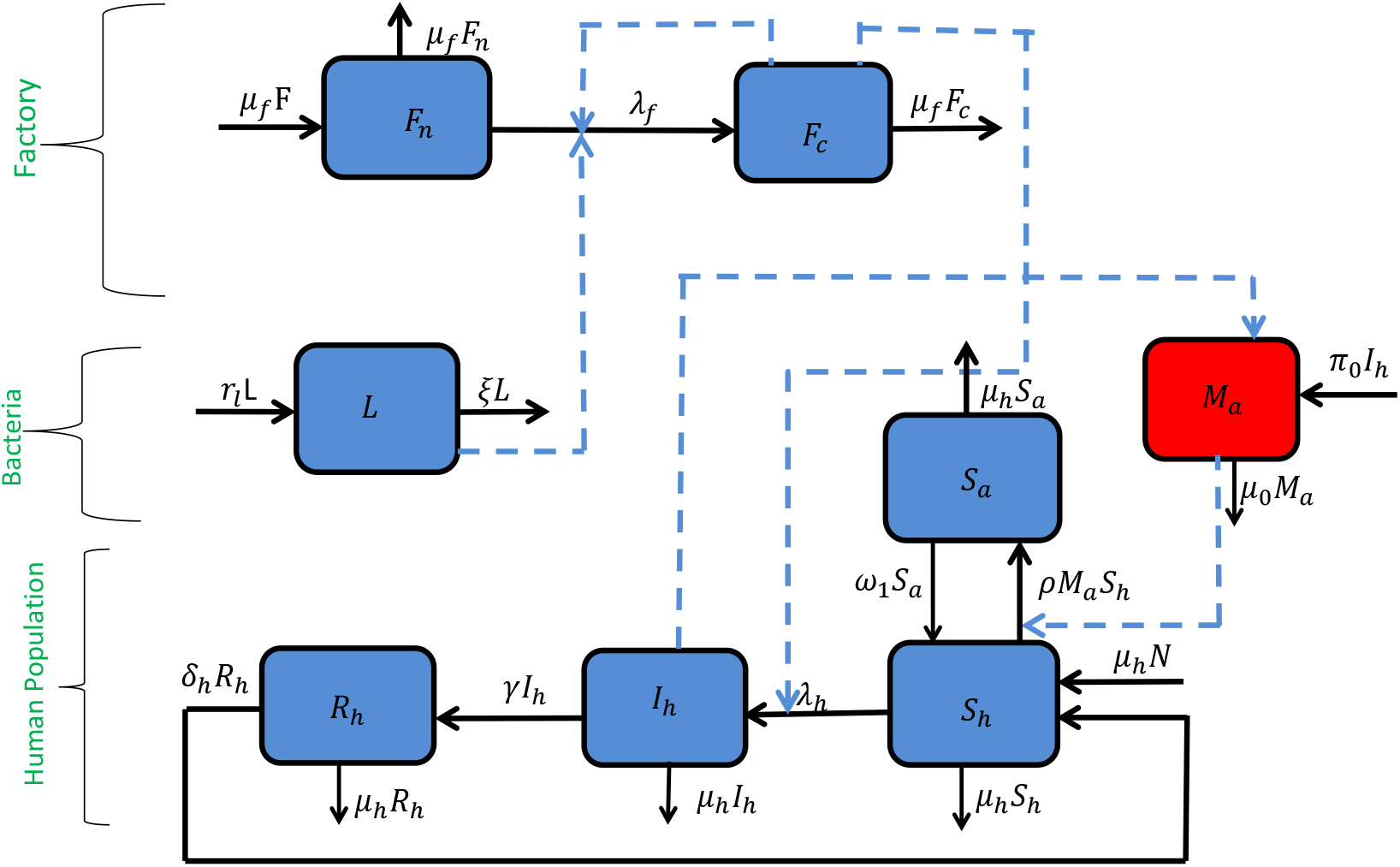
Shows the model flow diagram. The solid arrows represent the transitions between the compartments while the dotted lines represent the influences on the solid arrows and the compartments.

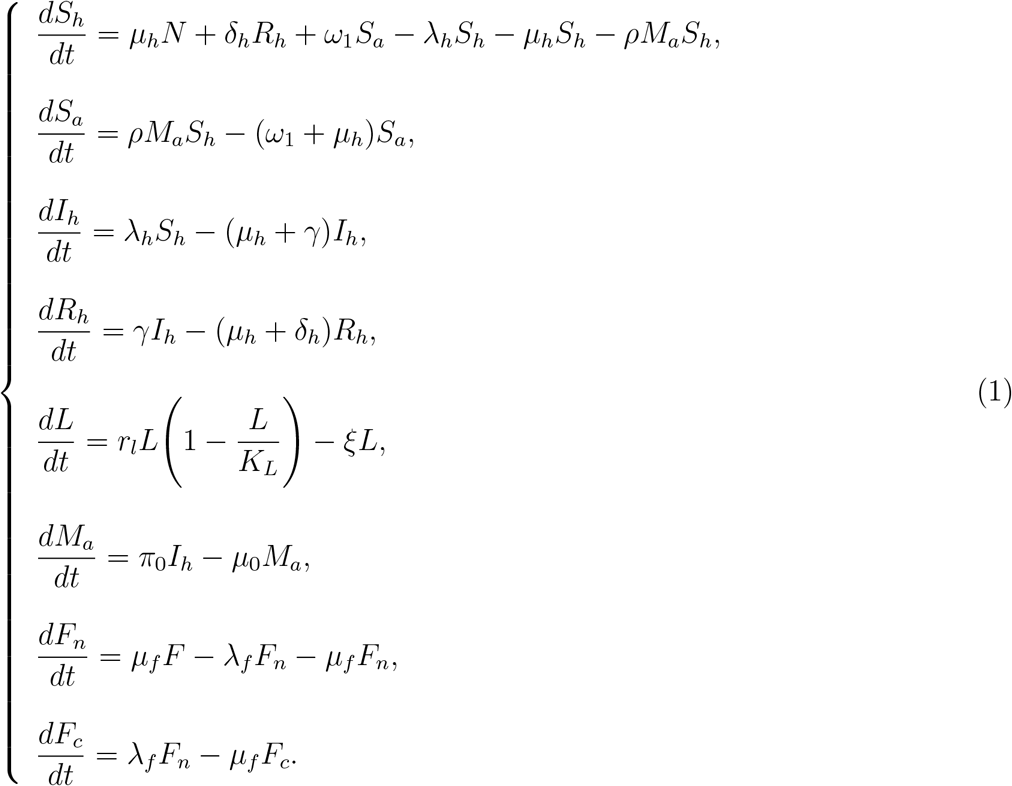

All parameters for the model system (1) are assumed to be non-negative for all time *t >* 0. By setting

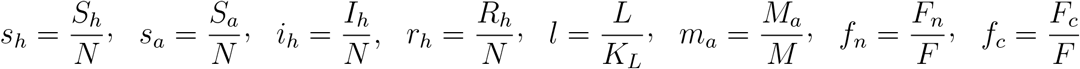

and given that *r*_*h*_(*t*) = 1 − *s*_*h*_(*t*) − *s*_*a*_(*t*) − *i*_*h*_(*t*) we have the following rescaled system

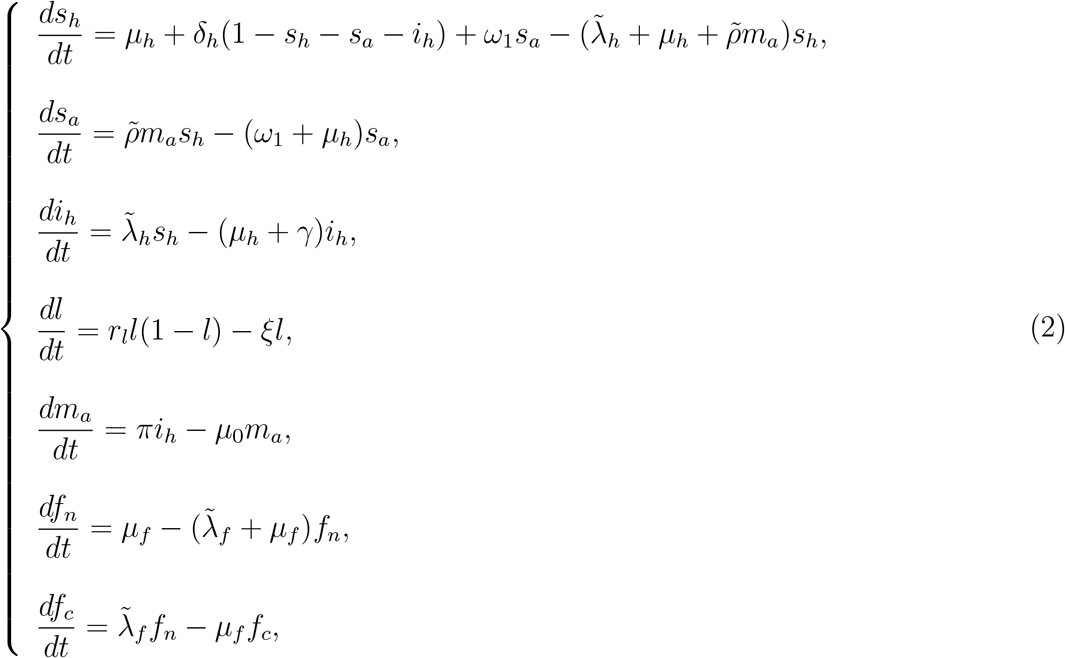

where

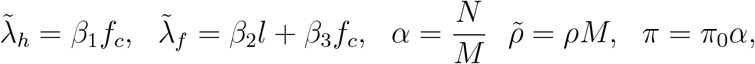

with 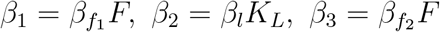 and initial conditions

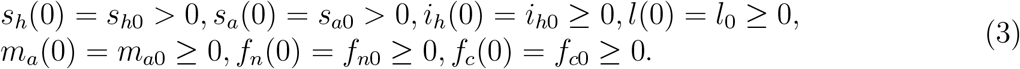

## 3 Model properties and analysis

### 3.1 Positivity of solutions

We prove the positivity of the solutions of model system (2) with initial conditions (3). First, we state the following Lemma as given in [23].

#### Lemma 1.

*Suppose* Ω ⊂ ℝ × *C*^*n*^ *is open, f*_*i*_ ∈ *C*(Ω, ℝ), *i* = 1, 2, …, *n. If*

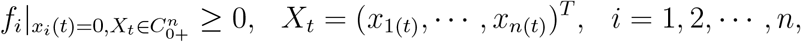

*then* 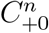 *is the invariant domain of the following equations*

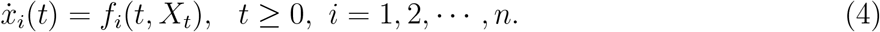

*If* 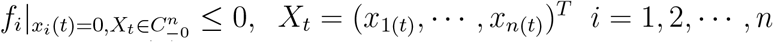, *then* 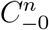 *is the invariant domain of equations (4)*.

We have the following Theorem on the invariance of system (2).

#### Theorem 1.

*Each solution* (*s*_*h*_(*t*), *s*_*a*_(*t*), *I*_*h*_(*t*), *l*(*t*), *m*_*a*_(*t*), *f*_*n*_(*t*), *f*_*c*_(*t*)) *of the model system (2) with the non-negative initial conditions (3) is non-negative for all t >* 0.

*Proof*. Let *X* = (*s*_*h*_, *s*_*a*_, *I*_*h*_, *l, m*_*a*_, *f*_*n*_, *f*_*c*_)^*T*^ and

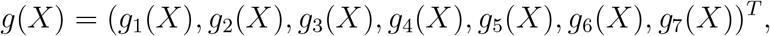

then we can re-write the model system (2) as follows:

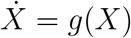

where

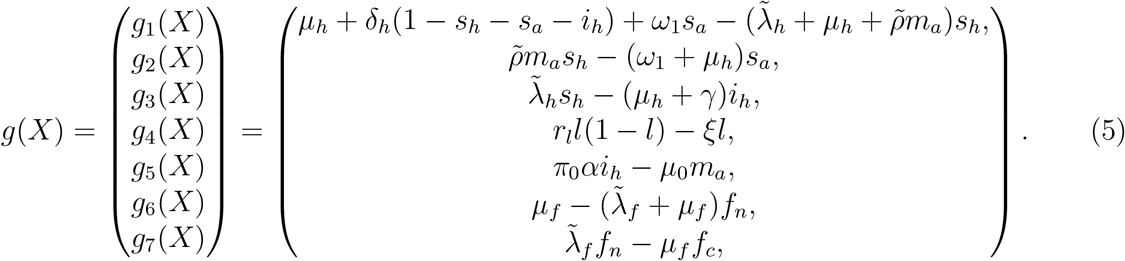

From (5), setting all the classes to zero, we have that

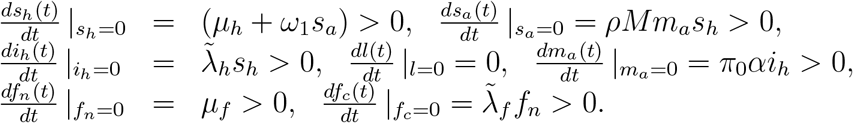

Thus, it follows that from Lemma 1 that 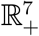 is an invariant set and positive. □

### 3.2 Existence and uniqueness of solutions

We now show that the solutions of systems (2) are bounded. We thus have the following result.

#### Theorem 2.

*The solutions of model system (2) are contained in the region* 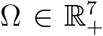, *which is given by* 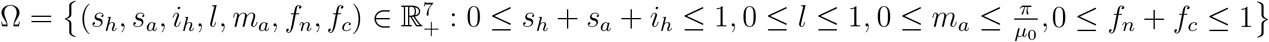 *for the initial conditions (3) in* Ω.

*Proof*. Considering the total change in the human population from the model system (2) given by

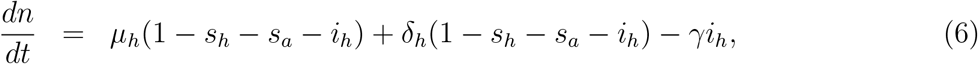

for *n* = *s*_*h*_ + *s*_*a*_ + *i*_*h*_ ≤ 1 we obtain

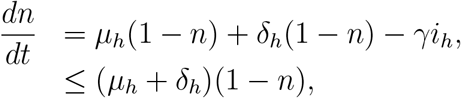

whose solution is

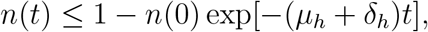

where *n*(0) = *s*_*h*_(0) + *s*_*a*_(0) + *i*_*h*_(0) is the initial condition. We note that 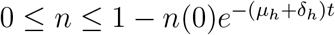, so that *n*(*t*) is bounded provided that *n*(0) ≥ 0.

The equation

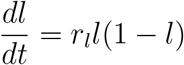

for the Listeria compartment has a standard solution for a logistic equation

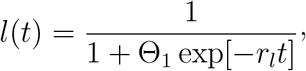

which is bounded with Θ_1_ = exp[−*c*], where *c* is a constant.

On the other hand, the total change in the amount of food products resulting from summing the last two equations of (2) is given by

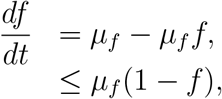

whose solution is

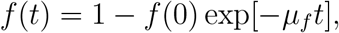

where *f* (0) = *f*_*n*_(0) + *f*_*c*_(0) Here, *f*_*c*_(*t*) ≤ 1 as *t → ∞* and hence it is bounded above. We thus, conclude that all the solutions of system (2) are bounded, biologically feasible and remains in Ω for all *t* ∈ [0, ∞). This completes the proof. □

### 3.3 Steady states and its analyses

The steady states of the model system (2) are obtained by equating the right side of equations (2) to zero, so that

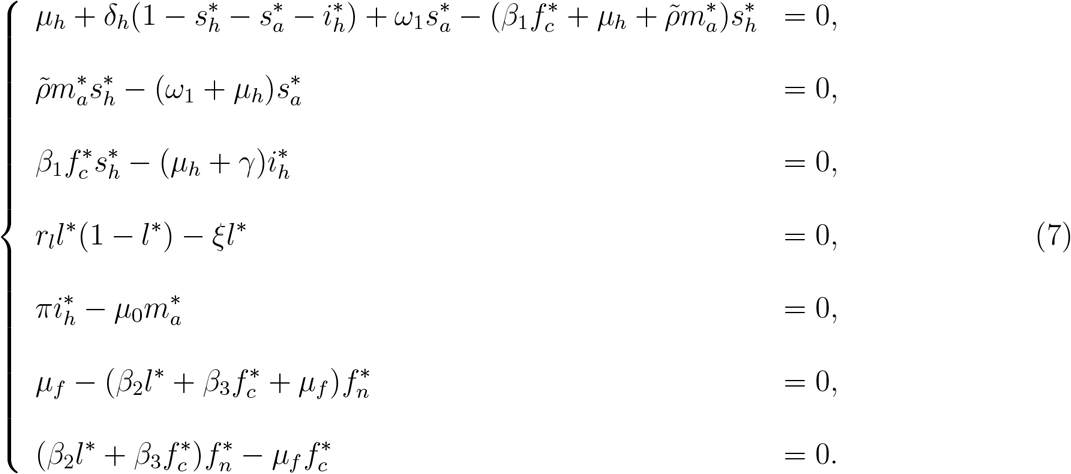

From the fourth equation of system (7), we have *l*^*^ = 0 or 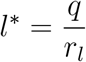, where *q* = (*r*_*l*_ — *ξ*) and *r*_*l*_ *> ξ*. We consider the two cases separately.

#### CASE A

If *l*^*^ = 0, (i.e. if there is no Listeria in the environment) then from the second last equation of (7) we have that

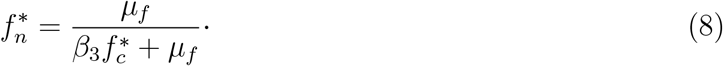

Substituting (8) into the last equation of (7) we obtain

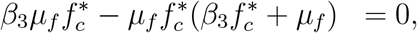

and upon simplification we have

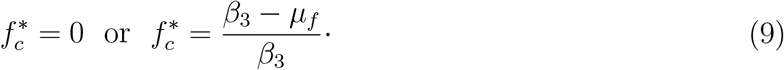

Thus, if *l*^*^ = 0 then 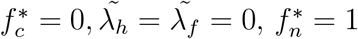 and 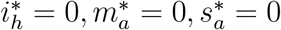. Also, from the first equation of (7)

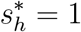

This results in the disease-free steady states (DFS) given by

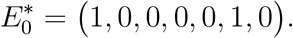

On the other hand, from (9), we have

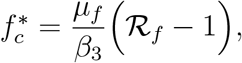

where

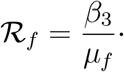

We thus have the following result on the existence of 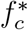.

##### Lemma 2.

*The existence of* 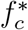 *is subject to* ℛ_*f*_ *>* 1.

However, we note that *β*_3_ is the contamination rate contributed by contaminated food products and 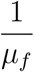 is the duration of food contamination. So, ℛ_*f*_ can be defined as the “food contamination threshold” that measures the growth of contaminated food due to the contamination of uncontaminated food products by contaminated food products. This is equivalent to the basic reproduction number (ℛ_0_) in disease modelling, see [24].

If 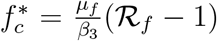, then expressing the second, third, and fifth equations of (7) in terms of 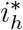 we obtain the following equation

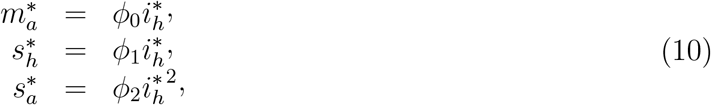

where 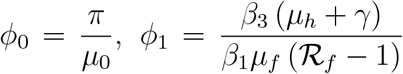 and 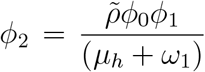.Substituting all the expressions in equation (10) into the first equation of (7) and after some algebraic simplifications we obtain the following quadratic equation where

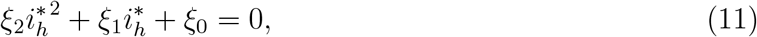

where

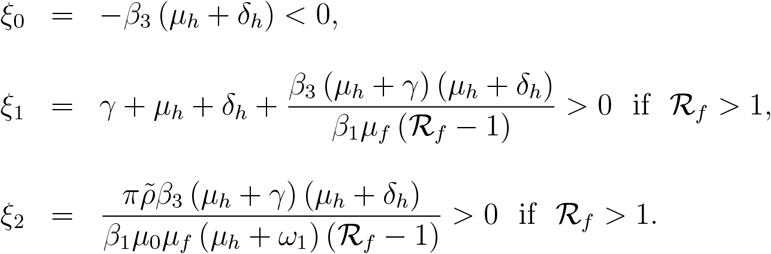

However, we note that the solutions of the quadratic equation (11) are given by

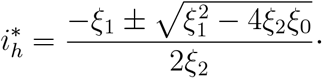

The solutions to (11) has one positive root when ℛ_*f*_ *>* 1. Biologically, this implies that the disease will persist and eventually invade the human population. This results in the Listeria steady state (LFS)

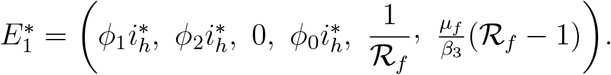

We note that at Listeria disease free steady state, there are contaminated food products which may result in Listeriosis infection in the human population.

##### Remark 1.

*We note that, when l*^*^ = 0, *we have two steady states* 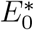 *and* 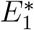. *The existence of* 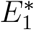 *is subject to the contaminated food generation number* (ℛ_*f*_) *been greater than 1. As long as ℛ*_*f*_ *>* 1, *even without Listeria in the environment, we will have the disease in the human population*.

#### CASE B

If 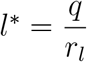, then from last equation of (7) solving for new 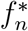, we have

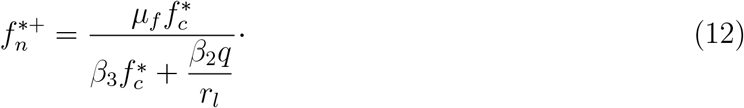

Substituting (12), into second last equation of (7) we obtain the following expression in terms of 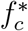 after some algebraic simplifications

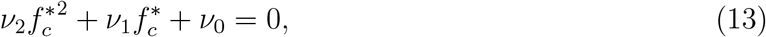

where

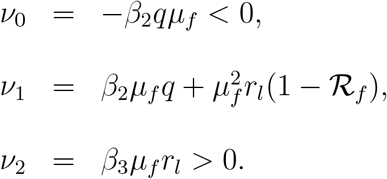

The solutions of the quadratic equation (13) given by

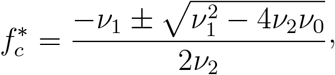

has one positive root irrespective of the signs of *ν*_1_. The solutions of 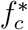 say 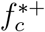 exists, but cannot be determined due to its intractability. Hence, as long as *l*^*^ = 1 we have a positive 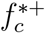.

Now, we express the second, third and fifth equation of (7) in terms of 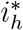 and obtain the following expressions

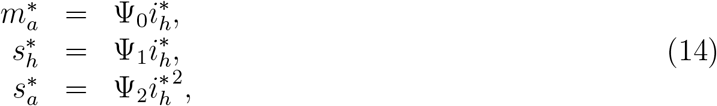

respectively, where 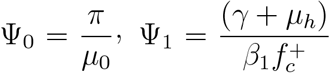 and 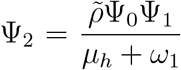 Similarly, substituting all the expressions from equations (14) into the first equation of (7) and after some algebraic manipulations we obtain the following quadratic equation in terms of 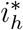

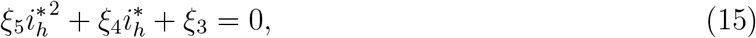

where

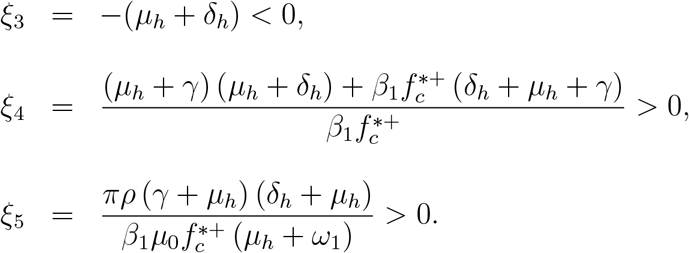

The solutions 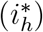 of the quadratic equation (15) given by

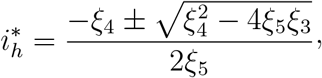

exists and has one positive root. We thus have the following result on the existence of new 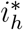. say 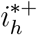.

##### Lemma 3.

*The steady state* 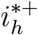 *exists whenever* 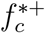 *exists*.

This results in the endemic steady states (ESS) given by

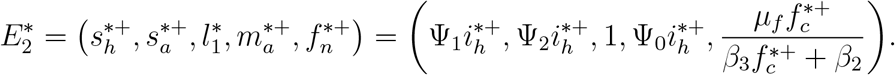

Hence, at endemic steady state, there are contaminated food products which result in the persistence of the Listeria infections in the human population.

#### 3.3.1 Local stability of the disease-free steady state

To analyse the local stability of the DFS, we show that the eigenvalues of the Jacobian matrix at DFS have negative real parts. We now state the following theorem for the DFS.

##### Theorem 3.

*The disease-free steady state* 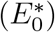 *is always stable whenever* ℛ_*f*_ *<* 1 *and r*_*l*_ *< ξ*.

*Proof*. The Jacobian of system (2) is given by the block matrix

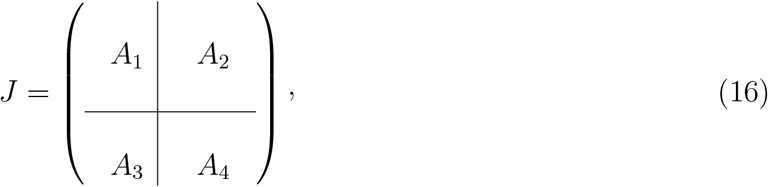

where

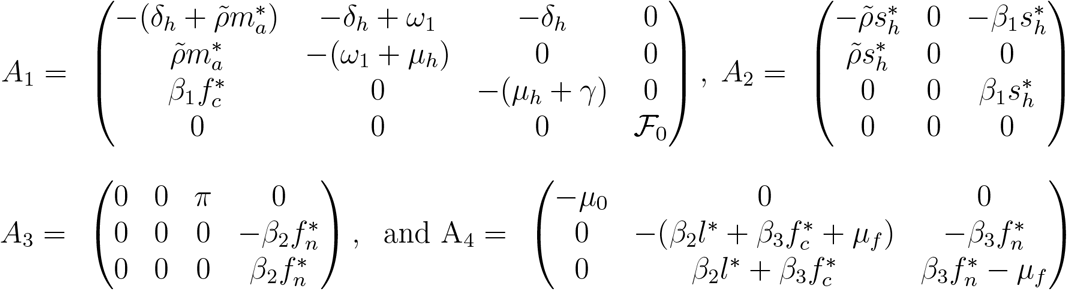

in which ℱ_0_ = *r*_*l*_ − *ξ* − 2*r*_*l*_*l*^*^. Evaluating (16) at DFS, we have that

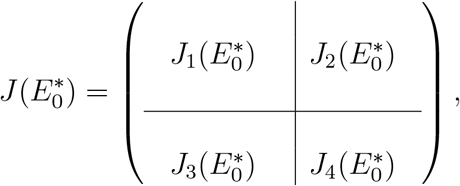

where

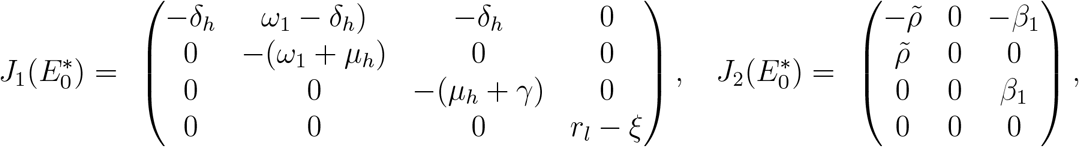

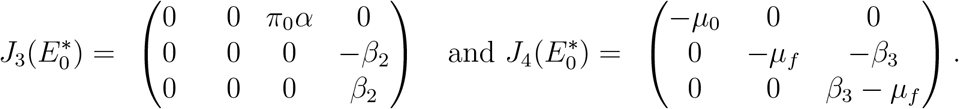

Similar to the eigenvalues approach [22], the eigenvalues of 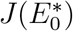 are: *λ*_1_ = −*δ*_*h*_, *λ*_2_ = −(*ω*_1_ + *µ*_*h*_), *λ*_3_ = −(*µ*_*h*_ + *γ*)*λ*_4_ = (*r*_*l*_ − *ξ*) *<* 0, if *r*_*l*_ *< ξ, λ*_5_ = −*µ*_0_, *λ*_6_ = −*µ*_*f*_ and *λ*_7_ = *µ*_*f*_ (ℛ _*f*_ − 1). We note eigenvalues are negatives hence 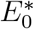 is stable. □

#### 3.3.2. Local Stability of the Listeria-free steady state

We state the following theorem for the local stability of Listeria-free steady state.

##### Theorem 4.

*The Listeria-free steady state* 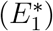 *is always stable whenever* ℛ_*f*_ *<* 1 *and r*_*l*_ *< ξ*.

*Proof*. Evaluating (16) at Listeria-free steady state, we have that

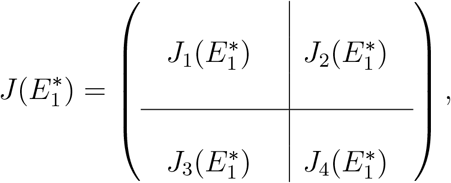

where

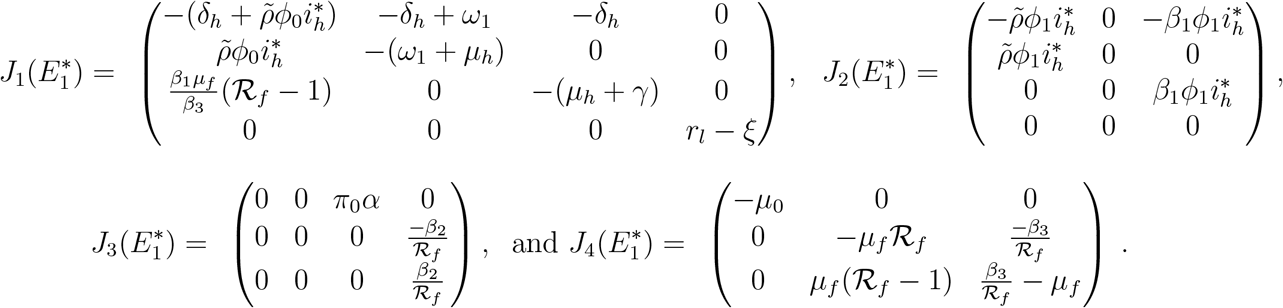

The eigenvalues from 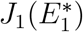 are: *λ*_1_ = (*r*_*l*_ − *ξ*) *<* 0 if *r*_*l*_ *< ξ* and the solutions to the cubic equation

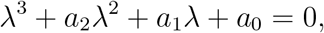

where

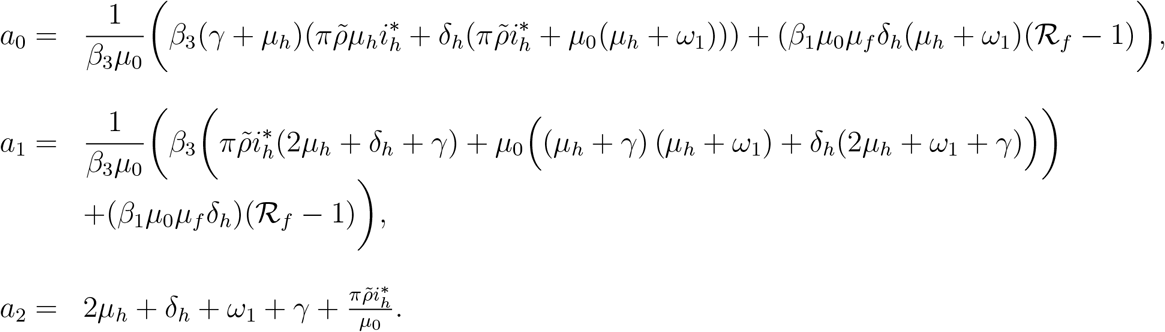

Given that *a*_0_, *a*_1_ and *a*_2_ are positive provided that ℛ_*f*_ *>* 1, we note that

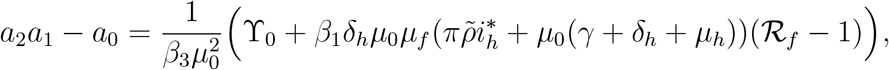

for

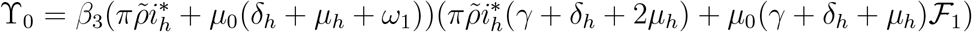

and ℱ_1_ = (2*µ*_*h*_ + *ω*_1_ + *γ*). We also, note that *a*_2_*a*_1_ − *a*_0_ *>* 0 if ℛ_*f*_ *>* 1. Hence, by the Routh-Hurwitz criterion the eigenvalues of 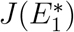 have negative real parts. The rest of the eigenvalues are determined from 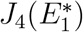, which are; *λ*_5_ = −*µ*_0_ and the solutions to the quadratic equation

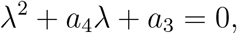

where

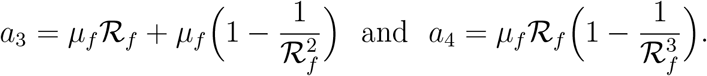

We thus have that *a*_3_ *>* 0 and *a*_4_ *>* 0 if ℛ_*f*_ *>* 1 and *a*_4_ is always positive. Therefore, the eigen-values have negative real parts by Routh-Hurwitz criteria. Hence, 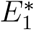 is locally asymptotically stable. □

#### 3.3.3 Local stability of the endemic steady state

We now state the following theorem on the local stability of endemic steady state.

##### Theorem 5.

*The endemic steady state* 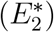, *is always locally asymptotically stable if* ℛ_*f*_ *>* 1 *and r*_*l*_ *> ξ*.

*Proof*. Evaluating (16) at endemic steady state, we have that

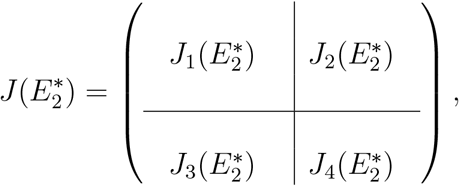

where

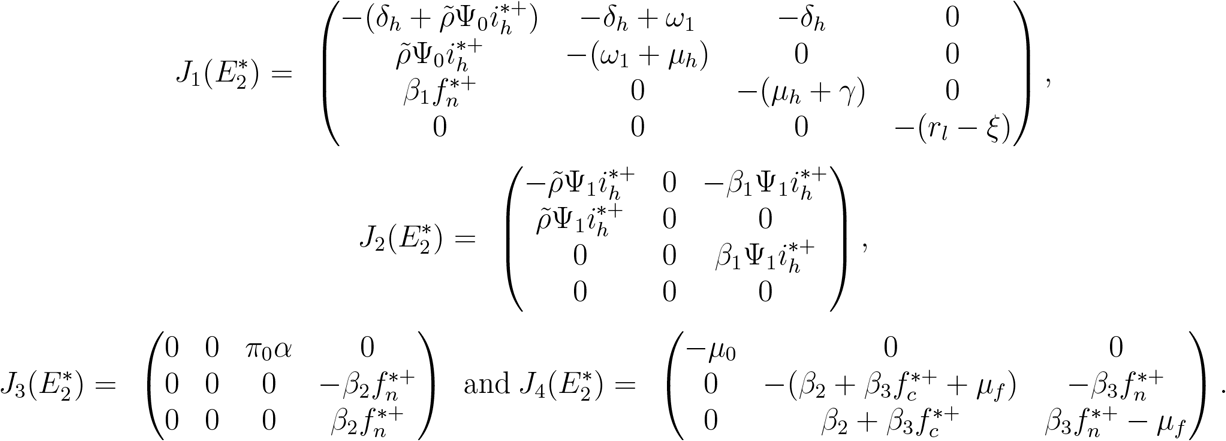

The eigenvalues from 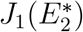 are: *λ*_1_ = − (*r*_*l*_ − *ξ*) *<* 0 if *r*_*l*_ *> ξ* and the solutions to the cubic equation

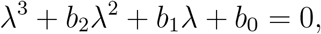

where

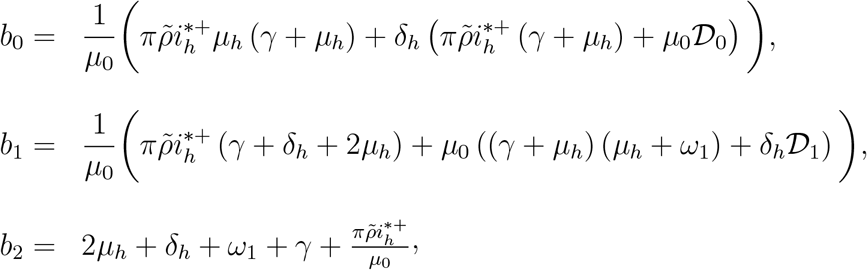

Note that *b*_0_, *b*_1_ and *b*_2_ are positive and

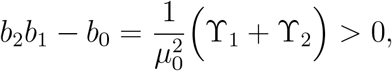

for

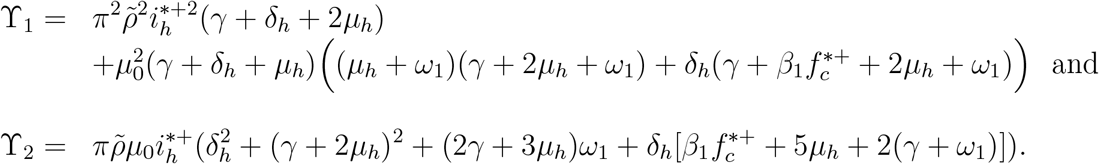

Thus, by the Routh-Hurwitz criteria, the eigenvalues have negative real parts. The remaining eigenvalues are determined from 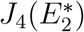, which are: *λ*_5_ = − *µ*_0_ and the solutions to the quadratic equation

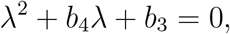

where

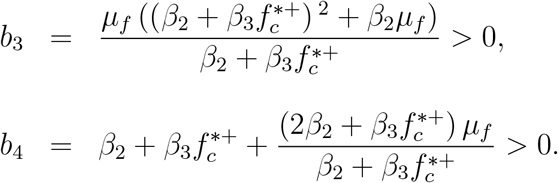

Hence, the eigenvalues have negative real parts by Routh-Hurwitz criterion. Therefore, all the eigenvalues of 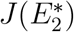 have negative real parts. Thus, 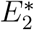 is locally asymptotically stable and otherwise unstable. □

## 4 Numerical results

### 4.1 Parameter values

Numerical simulations of the model system (2) were done using the set of parameters values given in Table 1. Most of the parameter values were estimated since there are very few mathematical models in literature, that have been done on *L. Monocytogenes* disease dynamics and hence the parameter values are elusive. We used a fourth order Runge-Kutta numerical scheme implemented in Matlab to perform the simulations with the initial conditions: *s*_*h*_(0) = 0.42, *s*_*a*_(0) = 0.53, *i*_*h*_(0) = 0.05, *l*(0) = 0.1, *m*_*a*_(0) = 0.25, *f*_*n*_ = 0.2 and *f*_*c*_ = 0.8. The initial conditions were hypothetically chosen for the numerical simulations presented in section and are thus only for illustrative purpose and do not represent any observed scenario.

**Table 1:** Parameter values used for numerical simulations.

| Parameter description | Symbol | Value (day<sup>-1</sup>) | Source |
| --- | --- | --- | --- |
| Mortality rate of humans | $\mu_h$ | 0.02/365 | [12] |
| Recovery rate of humans | $\gamma$ | 0.02 | Assumed |
| Rate of loss of immunity for humans | $\delta_h$ | 0.2 | [12] |
| Waning rate of aware susceptibles | $\omega_1$ | 0.25 | [15] |
| Rate of non-aware susceptibles to aware | $\tilde{\rho}$ | 0.9 | Assumed |
| Growth rate of Listeria | $r_l$ | 0.25 | [12] |
| Death rate of Listeria | $\xi$ | 0.056 | Assumed |
| Depletion rate of media campaigns | $\mu_0$ | 0.03 | [16] |
| Implementation rate of media campaigns | $\pi$ | 0.001 | [16] |
| Contact rate between Listeria and humans | $\beta_1$ | 0.0025 | Assumed |
| Food contamination rate by Listeria | $\beta_2$ | 0.09 | Assumed |
| Food contamination rate | $\beta_3$ | 0.0048 | Assumed |
| Removal rate of food products | $\mu_f$ | 0.056 | Assumed |

### 4.2 Sensitivity analysis

Sobol sensitivity analysis [25], were used to determine the model parameters that are sensitive to changes in some variable of the model system (2). We performed simulations for some chosen parameters *π, µ*_0_, 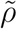, *ω*_1_, *β*_1_, *β*_2_, *β*_3_ and *µ*_*f*_ versus some of the model state variables *s*_*a*_, *i*_*h*_ and *f*_*c*_ to show their respective PRCCs over time. These parameters and state variables were selected because they are the most significant in the Listeria disease transmission and control according to our model formulation relating to the subject under investigation. The simulations were done using Matlab with 1000 runs over 800 days as depicted in Figures 4 and 3. The scatter plots for parameters with positive and negative PRCCs are also shown in Figure 2. Thus, an increase in the rate at which susceptible individuals move into the aware susceptible class results in a fewer number of humans been infected with the Listeriosis and parameter *ω*_1_ with negative correlation signifying that if aware individuals revert to being susceptible, then they are prone to contracting the disease as depicted in Figure 2 respectively. This highlights the importance of media campaigns in disease control.

**Figure 2:**
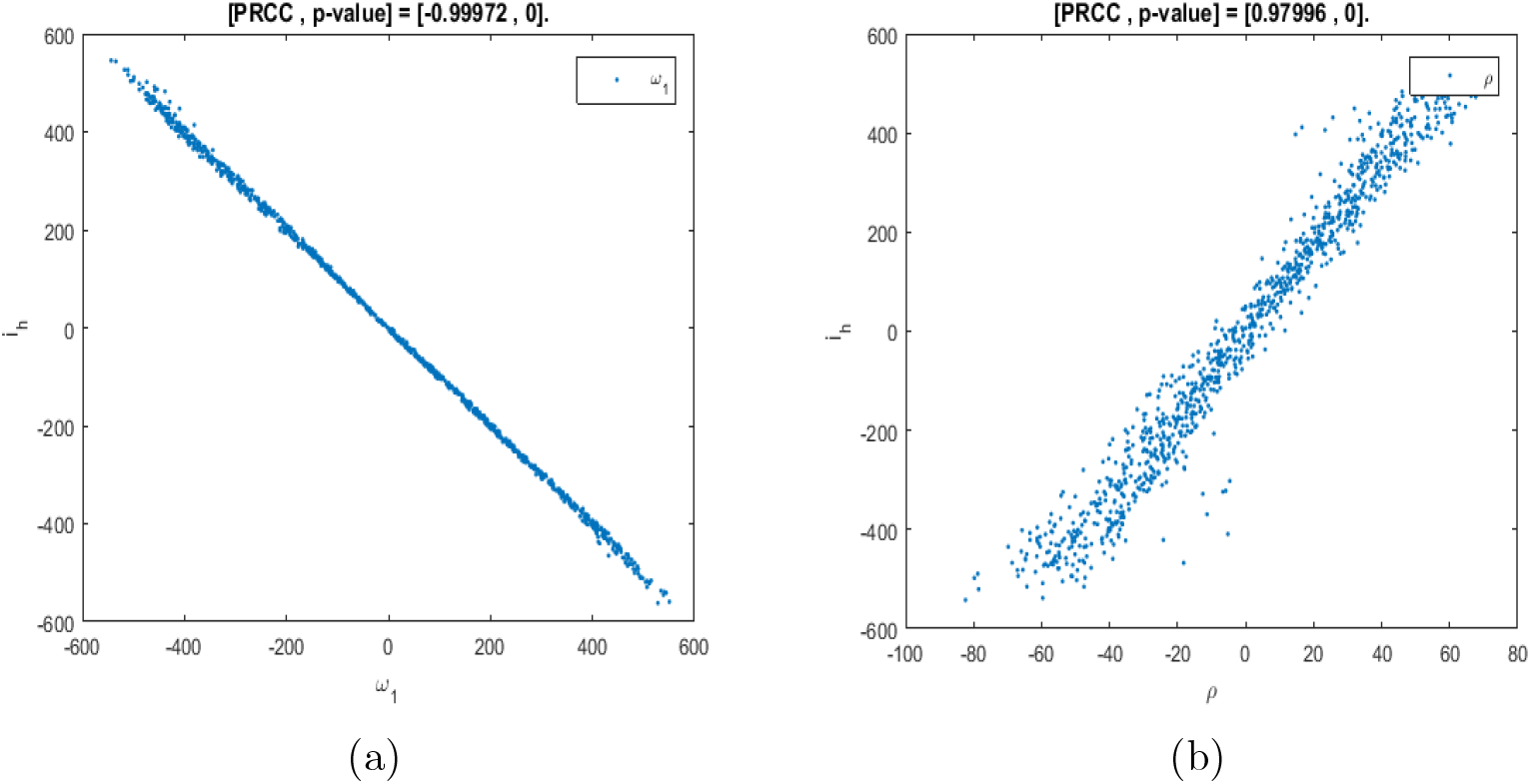
Scatter plots of parameters with the negative 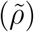 and positive (*ω*_1_) PRCCs.

**Figure 3:**
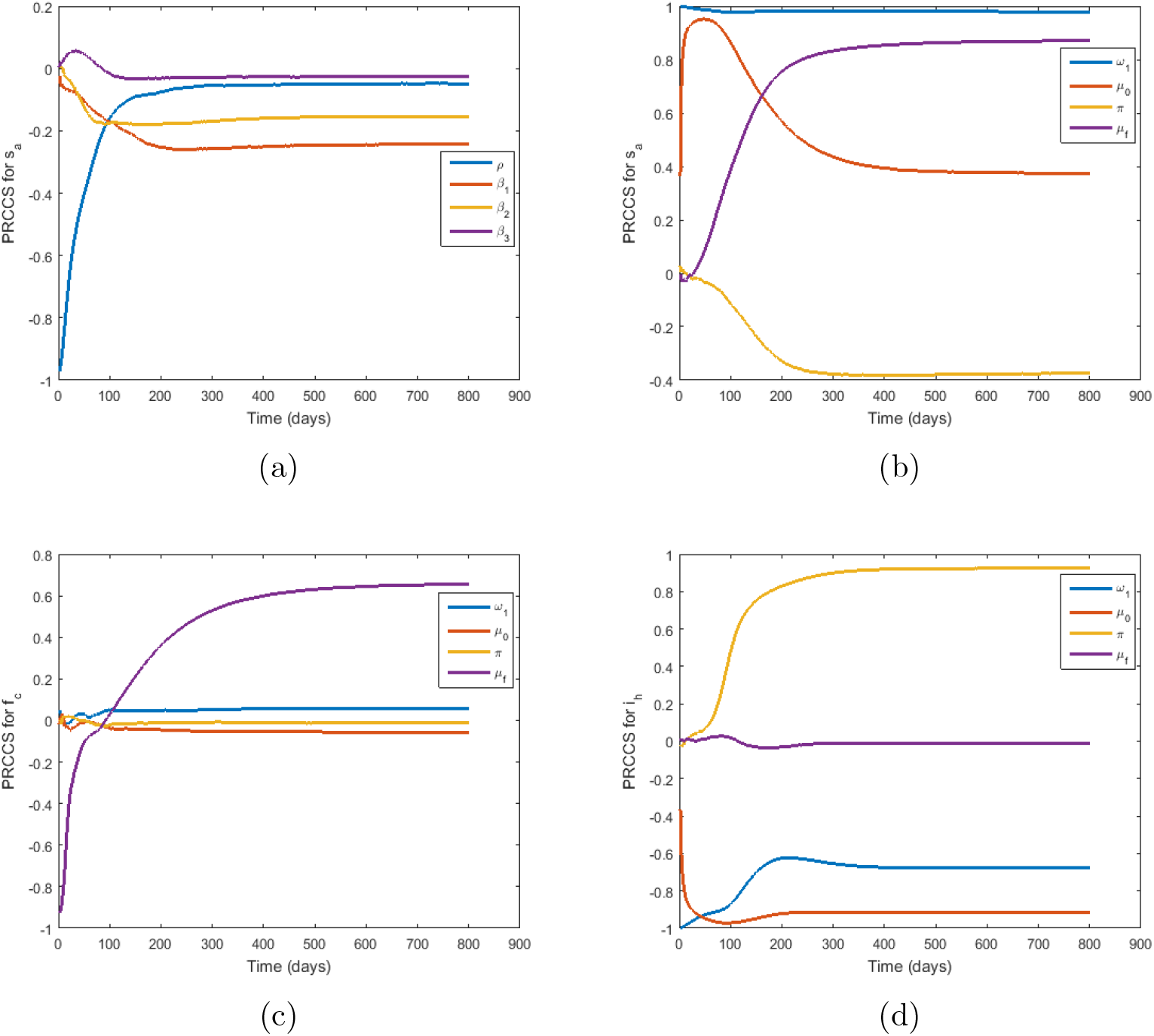
Partial correlation coefficients of parameters *π, µ*_0_, 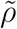, *ω*_1_, *β*_1_, *β*_2_, *β*_3_ and *µ*_*f*_ over time for state variables *s*_*a*_, *f*_*c*_ and *i*_*h*_ respectively. Note that, *ρ* in the legend represents 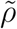 due to Matlab do not compile tilda.

**Figure 4:**
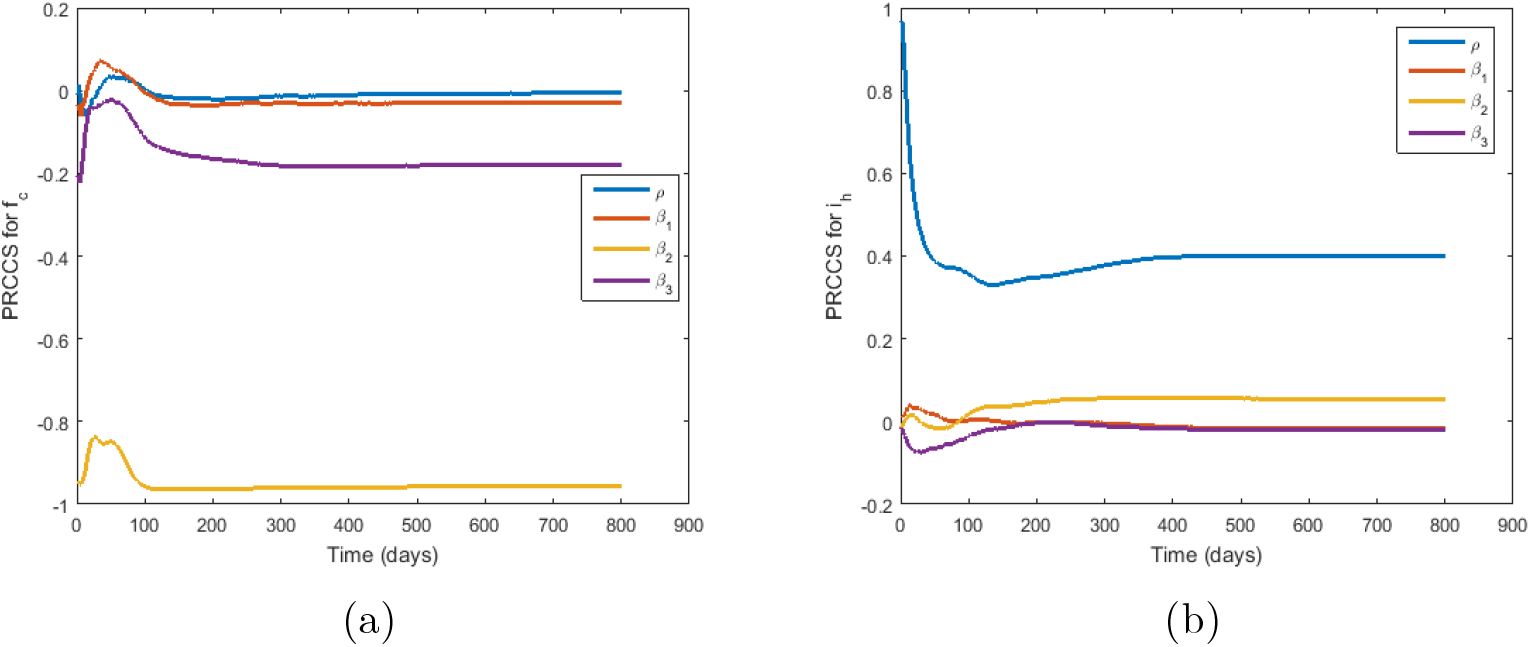
Partial correlation coefficients of parameters *π, µ*_0_, 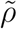, *ω*_1_, *β*_1_, *β*_2_ and *β*_3_ over time against *f*_*c*_ and *i*_*h*_ Note that, *ρ* in the legend represents 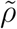 due to Matlab do not compile tilda.

### 4.3 Effects of varying *π* and *µ*_0_ on Listeriosis spread

This subsection is devoted to numerical simulations that show the effects of increasing and decreasing media campaigns over time. Figure 5(a) reveals that an increase in the awareness campaigns result in a decrease in the number of infected humans while a reduction of media campaigns results in more humans been infected with Listeriosis as shown in Figure 5(b).

**Figure 5:**
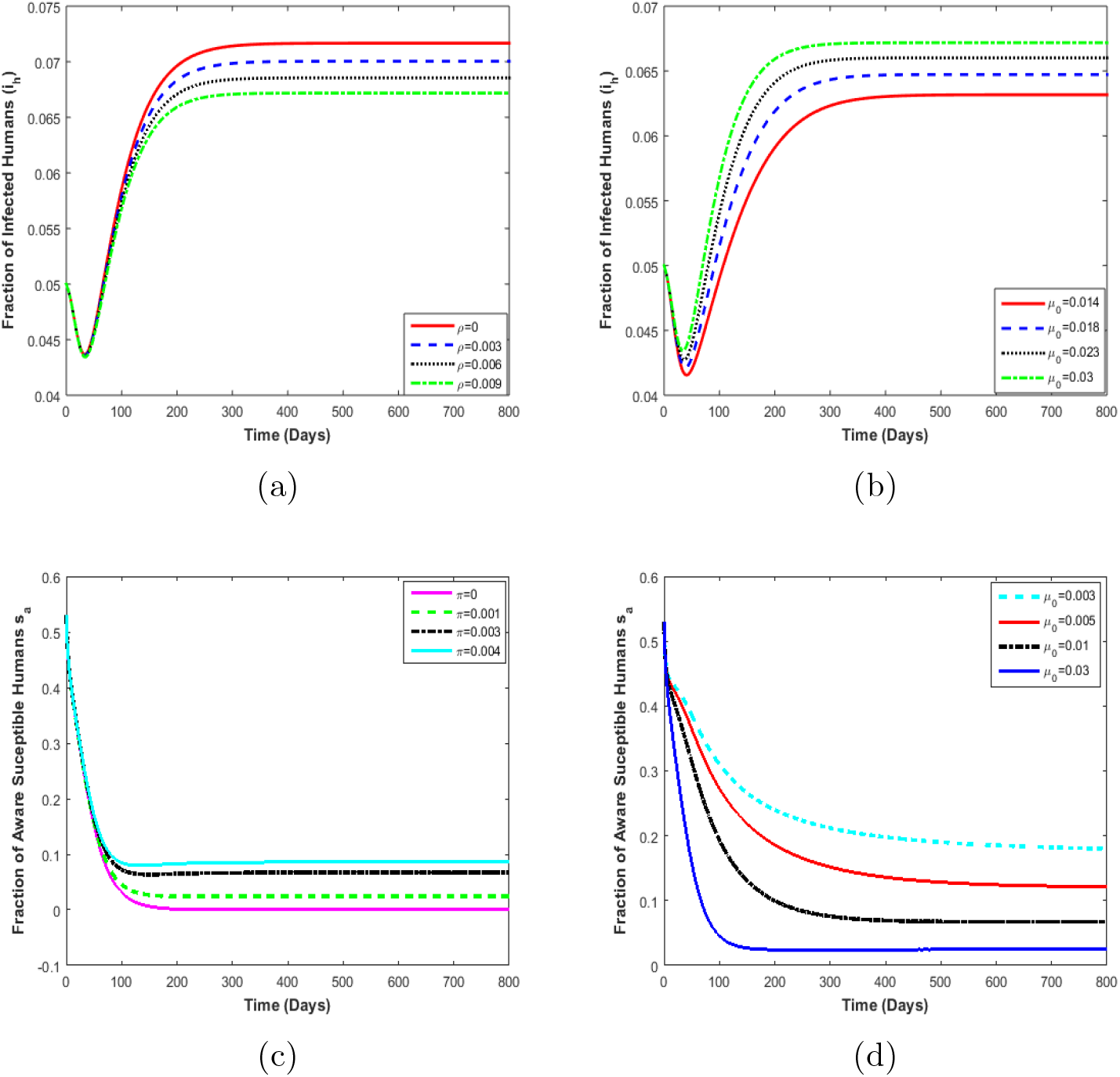
(a) Effects of implementing of media campaigns (*π*) on the fraction of infected humans. (b) Effects of depletion of media campaigns (*µ*_0_) on the fraction of infected humans. (c) Effects of implementation of media campaigns (*π*) on the fraction of aware susceptible humans. (d) Effects of depletion of media campaigns (*µ*_0_) on the fraction of aware susceptible humans.

### 4.4 Contour plot of *β*_3_ and *µ*_*f*_ on ℛ_*f*_

In Figure 6, we present a contour plot of the food contamination rate *β*_3_ and rate of food product removal, *µ*_*f*_, versus the food contamination constant, ℛ_*f*_. An increase in the contamination of non-contaminated food by contaminated food products results in an increase in the value of ℛ_*f*_. Hence, more humans get infected with Listeriosis. Also, an increase in the removal of contaminated food products results in a decrease in the values of ℛ_*f*_ which implies that fewer humans contract the disease. Note that, the changes in the value of *µ*_*f*_ do not significantly impact ℛ_*f*_ when compared to *β*_3_.

**Figure 6:**
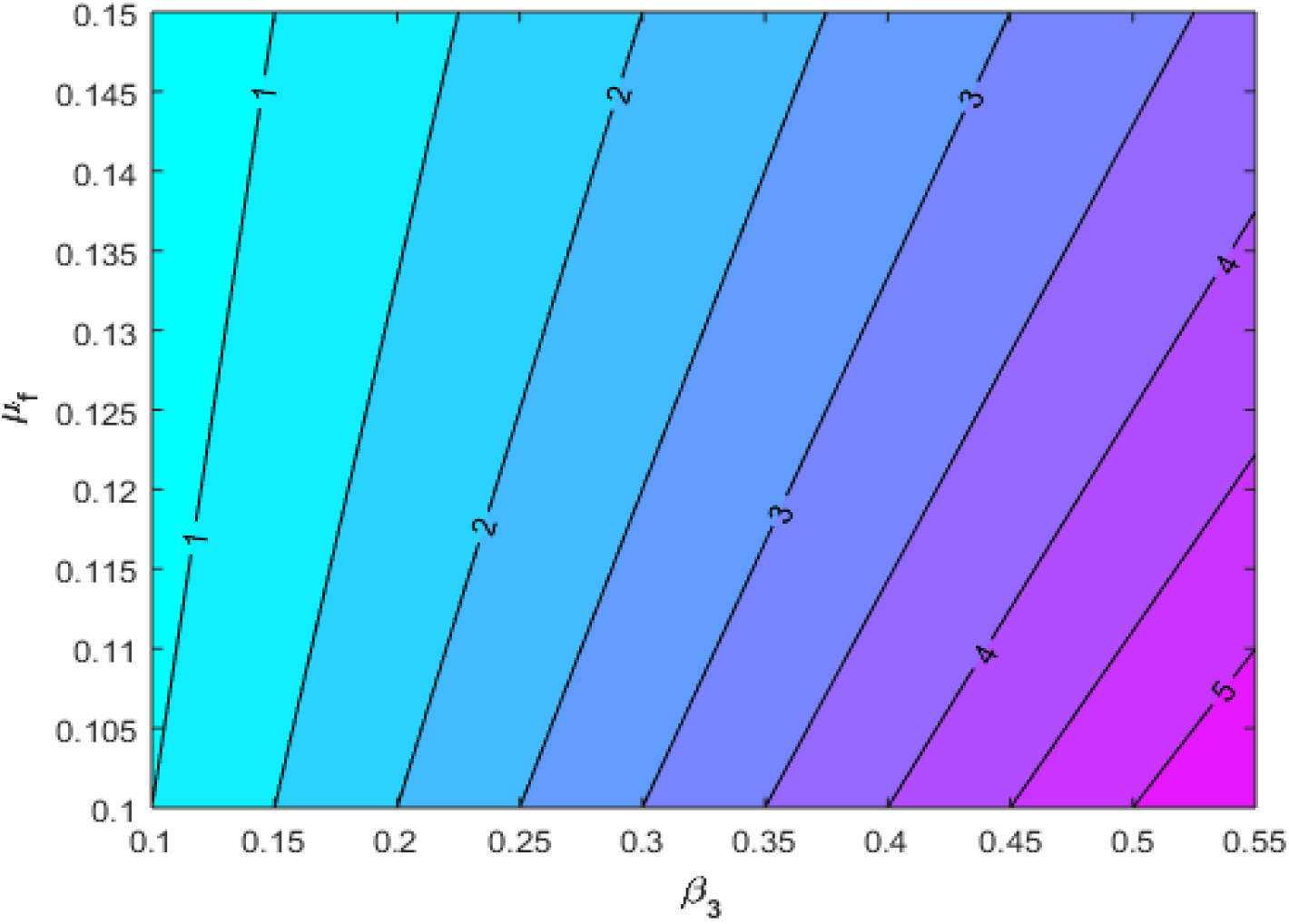
Contour plot of contaminated food generation number (ℛ_*f*_) as a function of *β*_3_ and *µ*_*f*_.

### 4.5 Effects of *β*_3_ and *µ*_*f*_ on ℛ_*f*_

Figure 7, illustrates a mesh plot showing the variation of *β*_3_, *µ*_*f*_ and the contaminated food generation number ℛ_*f*_. It is seen that an increase in the rate of food processing increases ℛ_*f*_ while an increase in the removal of contaminated food results in a decrease in ℛ_*f*_. Note that at the intersection of the plane ℛ_*f*_ = 1 and the mesh plot, we obtain all values of *β*_3_ and *µ*_*f*_ necessary for the eradication of the disease.

**Figure 7:**
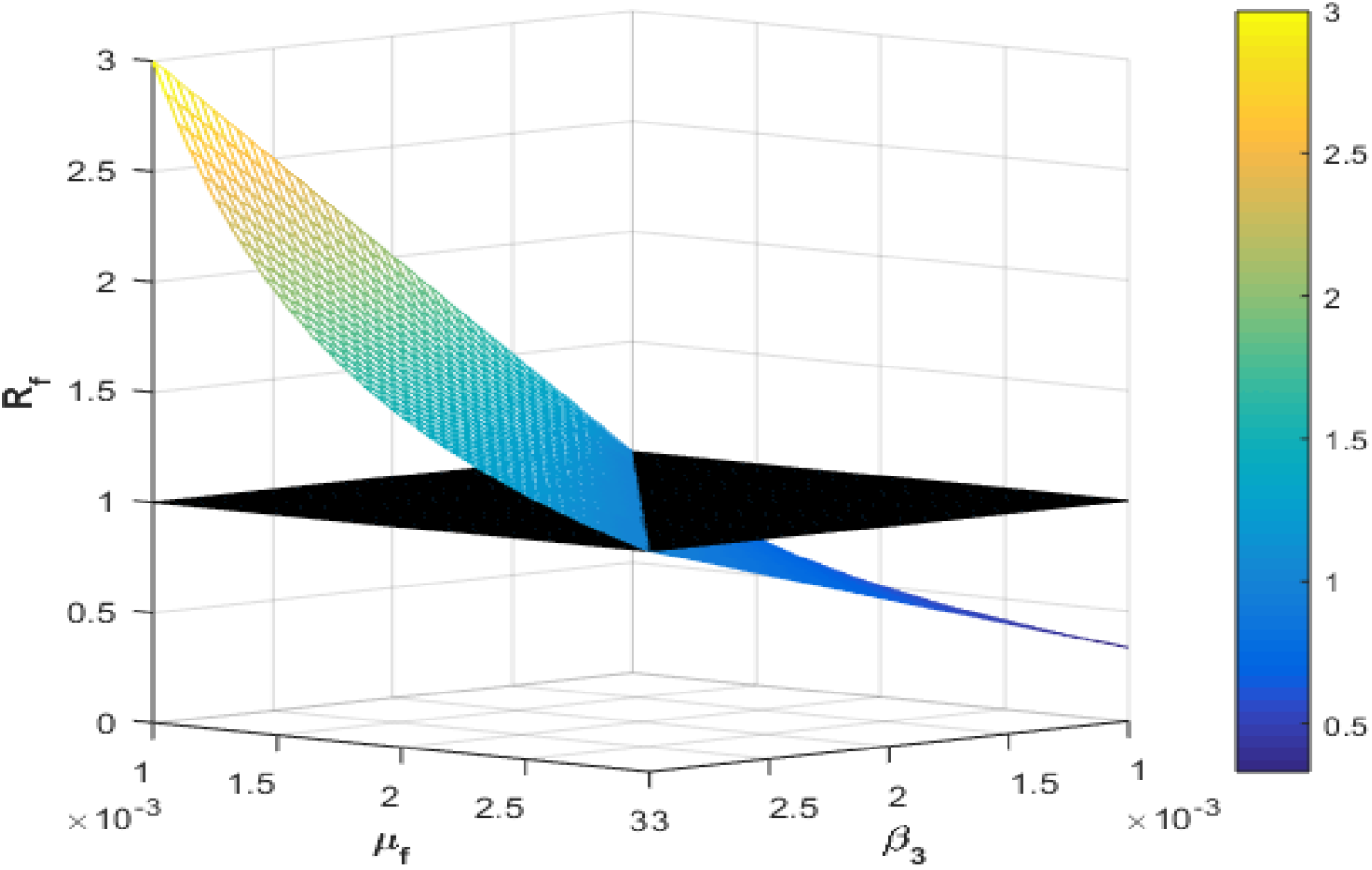
Effects of *β*_3_ and *µ*_*f*_ on the contamination food constant (ℛ_*f*_).

### 4.6 Impact of varying 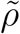 and *µ*_*f*_ on infected humans

Figure 8 depicts the effects of varying parameters 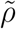 and *µ*_*f*_. In Figure 8(b), we observe that increasing 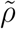 decreases the number of infected humans. This implies that media campaigns have the potential to reduce the number of infectives. On the other hand, an increase in 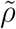 does not impact the value of the contaminated food generation number, ℛ_*f*_. On the other hand, increasing *µ*_*f*_ also reduces the number of infected individuals and ℛ_*f*_ (see Figure 8(a)). So the removal of contaminated food products during an outbreak is an important intervention in the control of Listeriosis.

**Figure 8:**
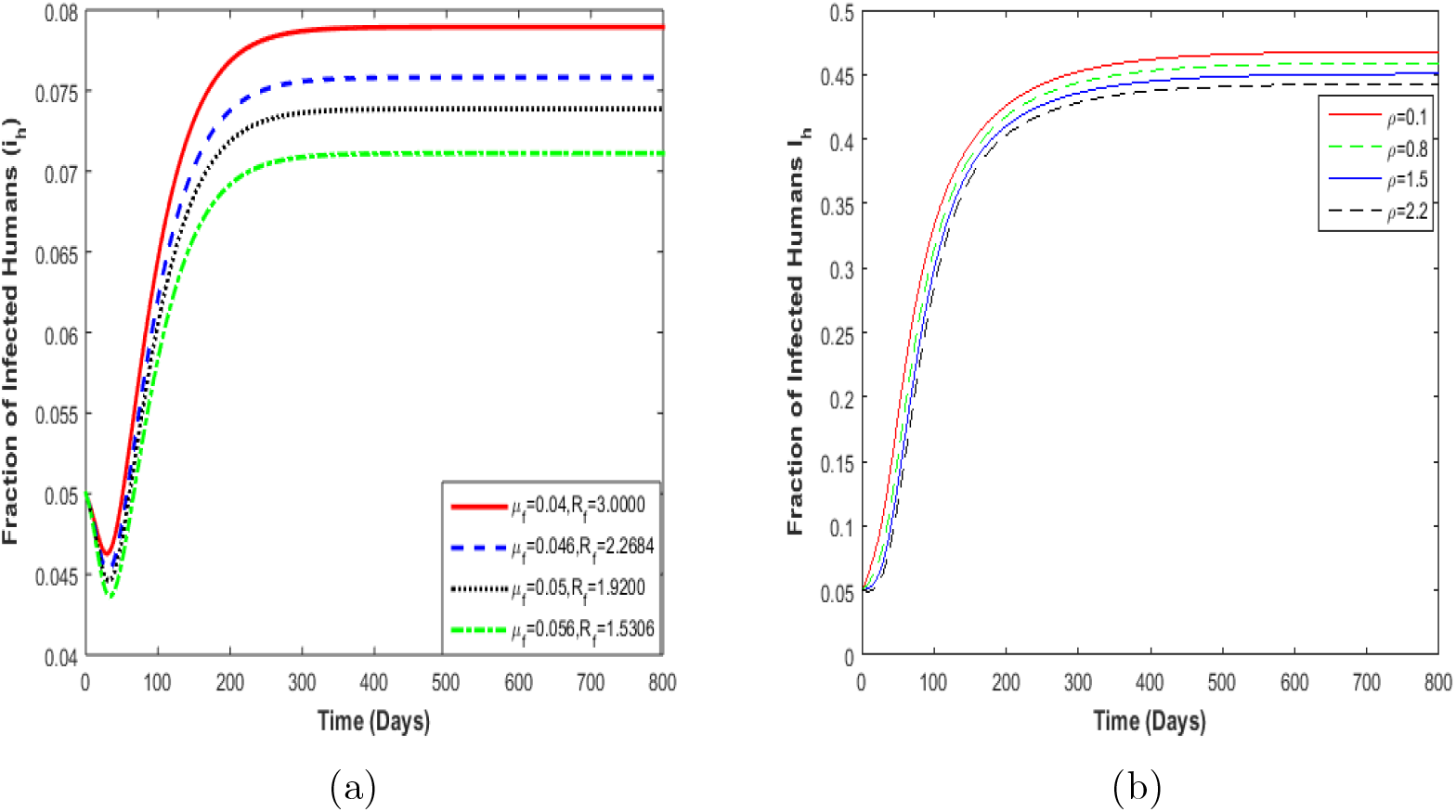
(a) Effects varying removal of contaminated food products (*µ*_*f*_) on the fraction of infected humans. (b) Effects varying the rate of non-aware susceptible becomes aware susceptible 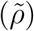 on the fraction of infected humans. The value for the ℛ_*f*_ is 1.5306.

## 5 Conclusion

A deterministic model on media campaigns’ potential role on Listeriosis disease transmissions was developed and analysed in this manuscript. Stability analyses of the model was done in terms of the food contamination constant ℛ_*f*_. The model exhibited three different steady states, which are: disease-free, Listeria-free and endemic equilibria. The disease-free and the Listeria-free steady states are locally asymptotically unstable if the net growth rate *r*_*l*_ *< ξ* and ℛ_*f*_ *<* 1 respectively, while the endemic equilibrium state is locally asymptotically stable if *r*_*l*_ *> ξ* and ℛ_*f*_ *>* 1

On the other hand from our numerical results, it was established that an increase in the removal of contaminated food products and increase in the rate at which the susceptible individuals (*s*_*h*_) becomes aware susceptible individuals (*s*_*a*_) leads to a decrease in the number of infected humans (see Figure 8). Thus to effectively control Listeria disease spread, policymakers, public health, governments and global stakeholders are advised to implement media campaigns that do not wane as time progresses. This means that media campaigns need to be effective. Further, these interventions from the campaigns should target mainly susceptible individuals on the danger of contracting Listeriosis. As people adhere to the media campaign effectively, it helps reduce and control the number of infected humans leading to less disease transmission.

The model presented in this paper is not without fallibility. The model was not fitted to epidemiological data and we assumed that the infectives do not interact with the aware susceptibles. During Listeriosis spread, the assumption has a negative impact on the human population since fewer un-aware individuals become aware of the disease. Despite these shortcomings, the results obtained in this paper are still implementable to help manage, control or contain Listeriosis disease transmission in the event of an outbreak.

## Data Availability

none

## Author’s contributions

C. W. Chukwu carried out the analytical, and numerical simulations. Both authors formulated and commented on the final version of this work.

## Acknowledgements

The authors would like to thank Faculty of Science in the University of Johannesburg for their support in the production of this manuscript. The authors are grateful to the anonymous reviewers for their lucrative suggestions and comments.

